# Trends in cardiac arrest mortality in the US, 1999-2020

**DOI:** 10.1101/2023.08.01.23293526

**Authors:** Karthik Gonuguntla, Muchi Ditah Chobufo, Ayesha Shaik, Neel Patel, Mouna Penmetsa, Harshith Thyagaturu, Amro Taha, Anas Alharbi, Yasar Sattar, Sudarshan Balla

## Abstract

**Importance:** Cardiac arrest affects over 600,000 in the US. Despite large-scale public health and educational initiatives, survival rates are lower in certain racial and socioeconomic groups.

**Objective:** To examine the trends in annual rates of cardiac arrests mortality among adults stratified by age, race, and gender in the U.S.

**Design:** A county-level cross-sectional longitudinal study using death data from the Centres for Disease Control and Prevention’s (CDC) Wide-Ranging Online Data for Epidemiologic Research (WONDER) database from 1999 to 2020.

**Setting:** Using the multiple causes of death dataset from the CDC’s WONDER database, cardiac arrests were identified using the International Classification of Diseases (ICD), tenth revision, clinical modification codes.

**Participants:** Individuals aged 15 years or more whose death was attributed to cardiac arrest.

**Exposures:** Calendar year.

**Main outcomes and measures:** National trends in annual mortality from cardiac arrest in select demographic groups and at the state level.

**Results:** Between 1999 and 2020, the annual cardiac arrest related AAMR (CA-MR) declined through 2019 (132.9 to 89.7 per 100,000 residents, followed by an increase in 2020 (104.5 per 100,000). White patients constituted 82% of all deaths and 51% were female. The overall CA-MR during the study period was 104.48 per 100,000 persons. The CA-MR was higher for men as compared with women (123.5 vs. 89.7 per 100,000) and higher for Black as compared with White adults (154.4 vs. 99.1 per 100,000).

**Conclusion and relevance:** CA-MR in the overall population has declined, followed by an increase in 2020, which is likely the impact of COVID-19 pandemic. There were also significant racial and sex differences in mortality rates.

**Visual Abstract:** Visual representation of cardiac arrest-related mortality and the impact of social vulnerability index on mortality rates^42^

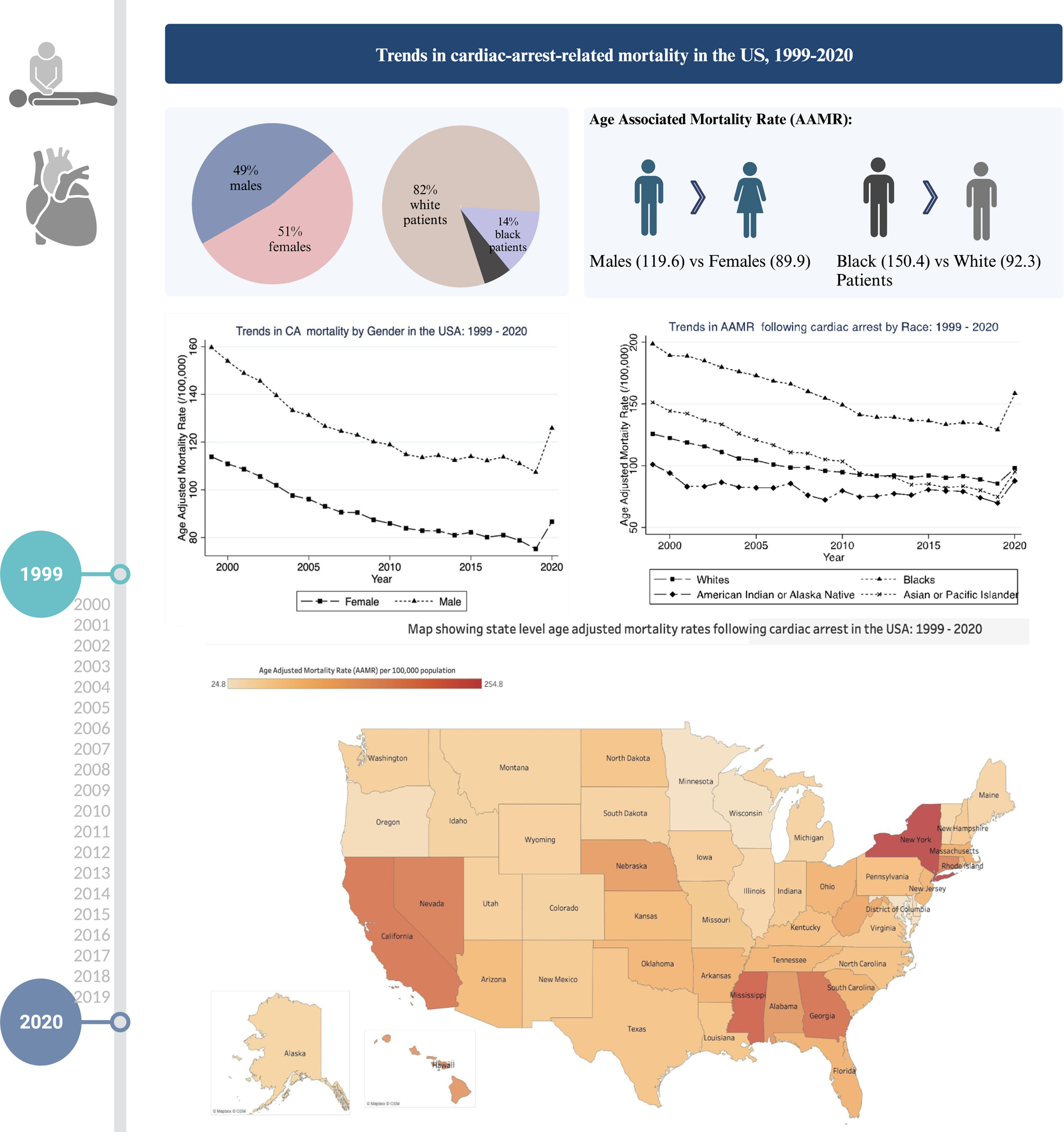

**Key Points:** Question

What are the temporal trends on cardiac arrest-related deaths in the United States (U.S.)?

Findings

In this national cross-sectional study spanning 22 years, there was an overall declining trend in cardiac arrest mortality across all age groups, gender, race, and regions. However, in 2020 there was an increase in the rates of cardiac arrest mortality, likely due to the COVID-19 pandemic. There was also a state-level variation in cardiac arrest mortality, with certain states showing an increase in mortality rates.

Meaning

There was an overall reduction in the age-adjusted mortality rates due to cardiac arrest. This decrease in mortality rate was consistent across all genders, races, and US regions. There was also significant racial and sex differences in mortality rates.

## Introduction

Cardiac arrest affects over 600,000 in the US^1,2^. The majority are related to an out-of-hospital cardiac arrest (OHCA) which contributes to over 340,000 events per year. Rates of in-hospital cardiac arrest (IHCA) are estimated to be around 292,000 events per year.^2^ Previous data have shown a steady decrease in the rates of IHCA, a similar decrease has not been noted in the rates of OHCA^2–5^ Our study extends the results of previously published papers as it incorporates all cardiac arrest death including IHCA and OHCA. We examined the incidence of cardiac arrest mortality per 1000 persons, rather than looking at the case survival rates among those with cardiac arrest. Despite large-scale public health and educational initiatives to train the public to recognize and perform bystander cardiopulmonary resuscitation (CPR), variation in survival rates still exists among racial and socioeconomic groups ^6^. It has been reported that the likelihood of survival drops by 7-10% for each minute without CPR, only 25% of OHCA receive bystander CPR, and less than 3% of the U.S. public receives annual CPR training ^7^.

Both IHCA and OHCA demonstrate significant overlap in relation to baseline co-morbidities, sex/racial and regional differences^8^. Various changes have been made to the American Heart Association resuscitation guidelines regarding the skills and ability to manage cardiac arrests such as rapid response teams to better manage and standardize CPR, intensive care monitoring, hypothermia protocol, and advanced mechanical circulatory support devices ^9^. Prior studies have reported sex- and racial differences in survival at the patient- and neighborhood level from OHCA and IHC^10^. Racial disparities also play a key role in survival rate variations as the racial composition of a community has been shown to influence the likelihood that an individual with an OHCA survives to hospital discharge or has favorable neurological survival^11–13^. Cardiac arrests in a majority of African American communities were less likely to survive discharge compared to the White population^14^. The purpose of our study was to assess the trends of cardiac arrest mortality in the US over 22 years at a population level.

## Methods

The Centers for Disease Control and Prevention’s Wide-Ranging Online Data for Epidemiologic Research (CDC WONDER) multiple-cause-of-death database was utilized to obtain deaths occurring in the US with cardiac arrest listed as one of the causes of death estimates from January 1999 to December 2020^11,15,16^. We studied cardiac arrests as a contributing or underlying cause of death on nationwide death certificates using the International Statistical Classification of Diseases, Tenth Revision, (ICD-10) code I46 (I46.0, I46.1, I46.9). This data is based on U.S. residents’ death certificates, and each death certificate contains a single underlying cause of death, up to 20 additional multiple causes, and sociodemographic data. The underlying cause of death is defined as the “disease or injury which initiated the train of events leading directly to death.” When >1 cause or condition is listed on the death certificate, the underlying cause is determined by the sequence of conditions on the certificate, provisions of the ICD, and associated selection rules and modifications. The study was exempt from our institutional review board approval, given the deidentified nature of the database.

Age-adjusted mortality rate (AAMR) and corresponding standard errors for all deaths with cardiac arrests listed as the underlying cause of death were obtained from 1999 to 2020 for the entire U.S. We further stratified the AAMR by age, sex, race/ethnicity, census region, state, and urban-rural classification. Race was categorized as White patients, African American/Black patients, American Indians/Alaska Natives, and Asian/Pacific Islanders. Ethnicity was categorized as Hispanic and non-Hispanic. As mentioned on the death certificates, race, and ethnicity were assessed in accordance with standards from the US Office of Management and Budget.

### Statistical analysis

Cardiac arrest-related age-adjusted mortality rates (CA-MR) per 100 000 persons were determined. AAMRs, standard errors (SE), and 95% confidence intervals (CI) were calculated by standardizing CA-related deaths to the corresponding year 2000 U.S population, as previously described. The Joinpoint regression program (version 4.9.1.0; National Cancer Institute) was used to describe trends in CA-MR^17^. Temporal trends in AAMR were determined by fitting log-linear regression models. We applied Joinpoint segmented regression to identify inflection points in the temporal trends of CA-MR from 1999 to 2020 based on published methodological guidelines^18^. Data points with less than 20 mortality events were flagged unreliable and treated as missing data. For data containing 22-26 time points, the guidelines recommend that the analysis identifies a maximum of 4 joinpoints across the study period. In the current investigation, 22 years were included; therefore, the Joinpoint regression statistical software was set to determine a maximum of 4 joinpoints where significant temporal variation existed in the trend. Best fit models with corresponding joinpoints were suggested by the software. Therefore, zero to a maximum of 4 joinpoints were allowed to be identified. The Grid Search method (2, 2, 0), permutation test, and parametric method were used to estimate average annual percent change (APC) and 95% confidence intervals (CIs). The average annual percent change over 22 years was then calculated using a weighted average of the slope coefficients of the underlying joinpoint regression line with weights equal to the length of each segment divided by 22. AAPC was considered increasing or decreasing if the slope describing the change in age-adjusted mortality was significantly different from 0 using 2-tailed t testing with p values <0.05 considered statistically significant.

## Results

The CA related deaths between 1999-2020 were 7,710,211 in the entire US. Of these, 49% were males, and 51% were women. The majority were White patients (82%), and Black patients constituted 14% of the overall cohort. The overall CA-MR during the study duration was 104.48 per 100 000.

AAMR was higher for males compared with females (123.49 per 100,000 persons vs 89.66 per 100,000 persons) and higher for Black adults compared to White patients (154.41 per 100,000 persons vs 99.13 per 100,000 persons, respectively). Upon further stratifying race and gender, AAMR was highest in Black males (182.16 per 100,000 persons) followed by Black females (135.22 per 100,000 persons), White males (117.65 per 100,000 persons), and White females (80.78 per 100,000 persons) respectively. The overall CA-MR decreased from 132.86 per 100,000 persons in 1999 to 89.67 per 100,000 persons in 2019 followed by an increase in 2020 to 104.48 per 100,000 persons. AAMR in all races and sex has trended down from 1999 to 2019 with an increase in AAMR in 2020 (Figure 1a, 1b). Among White females, AAMR decreased from 1999 to 2007 (APC, −3.3% [95% CI, −3.7 to −2.9]) and 2007 to 2018 (APC, −0.9% [95% CI, −1.1 to 0.6]) with a significant increase from 2018 to 2020 (APC, 4.7% [95% CI, 1.1 to 8.4]), overall (APC, −1.3% [95% CI, −1.7 to 0.9]). Among Black females, AAMR decreased from 1999 to 2018 (APC, −2.4% [95% CI, −2.6 to −2.3]) followed by a rise between 2018 and 2020 (APC, 8.7% [95% CI, 3.2-14.5]). Among Black males, AAMR decreased from 1999 to 2018 (APC, −2.2% [95% CI, −2.4 to −2.0]) followed by a rise between 2018 and 2020 (APC, 11.7% [95% CI, 4.3-19.7]). Among White males, there was a steady decrease in the AAMR from 1999 to 2009 (APC, −2.8% [95% CI, −3.4 to −2.3]) followed by another slow decline from 2009 and 2020 (APC, −0.6% [95% CI, −1.1to −0.1]).

**Figure 1a:**
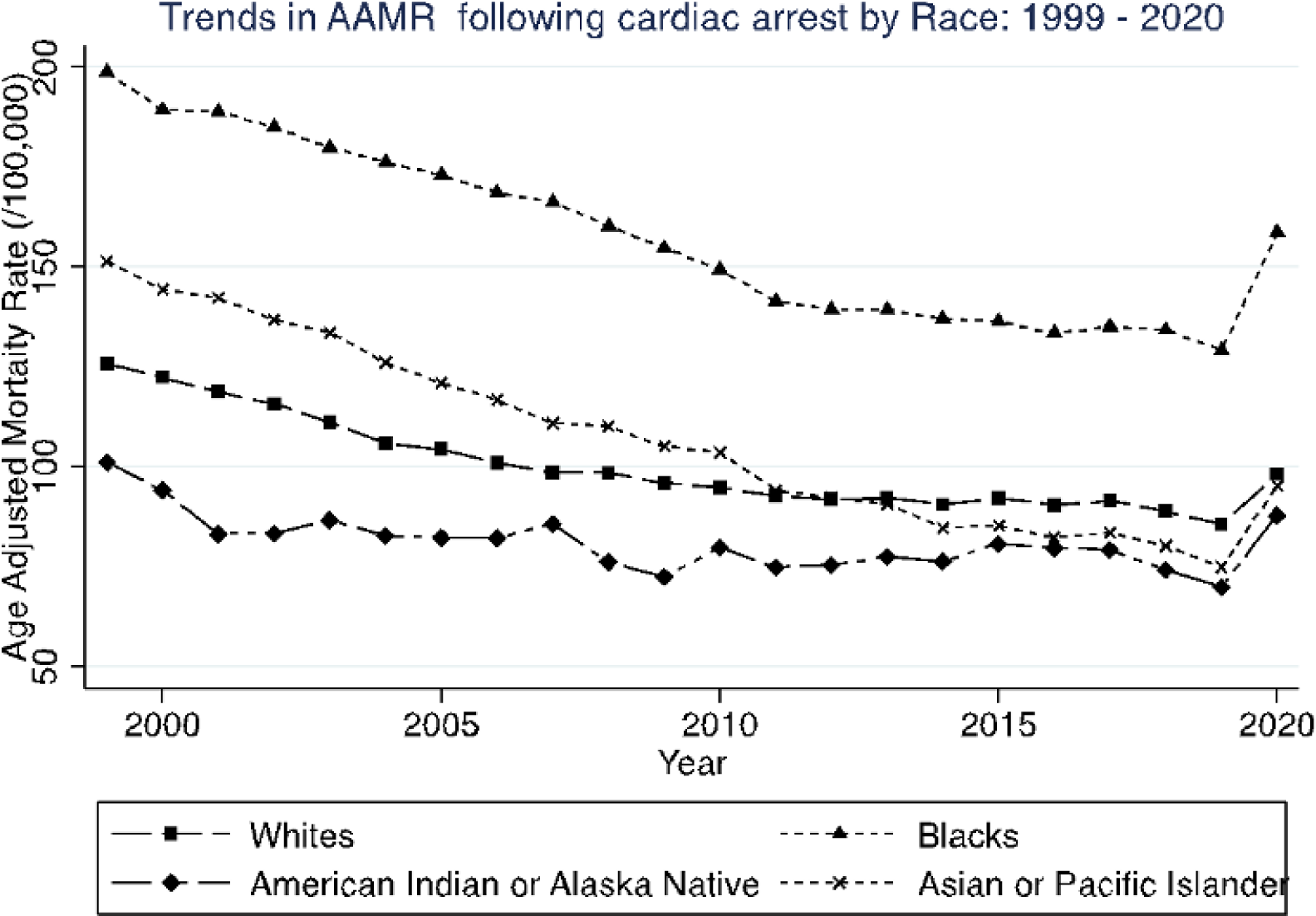
Trends in cardiac arrests related mortality by race.

**Figure 1b:**
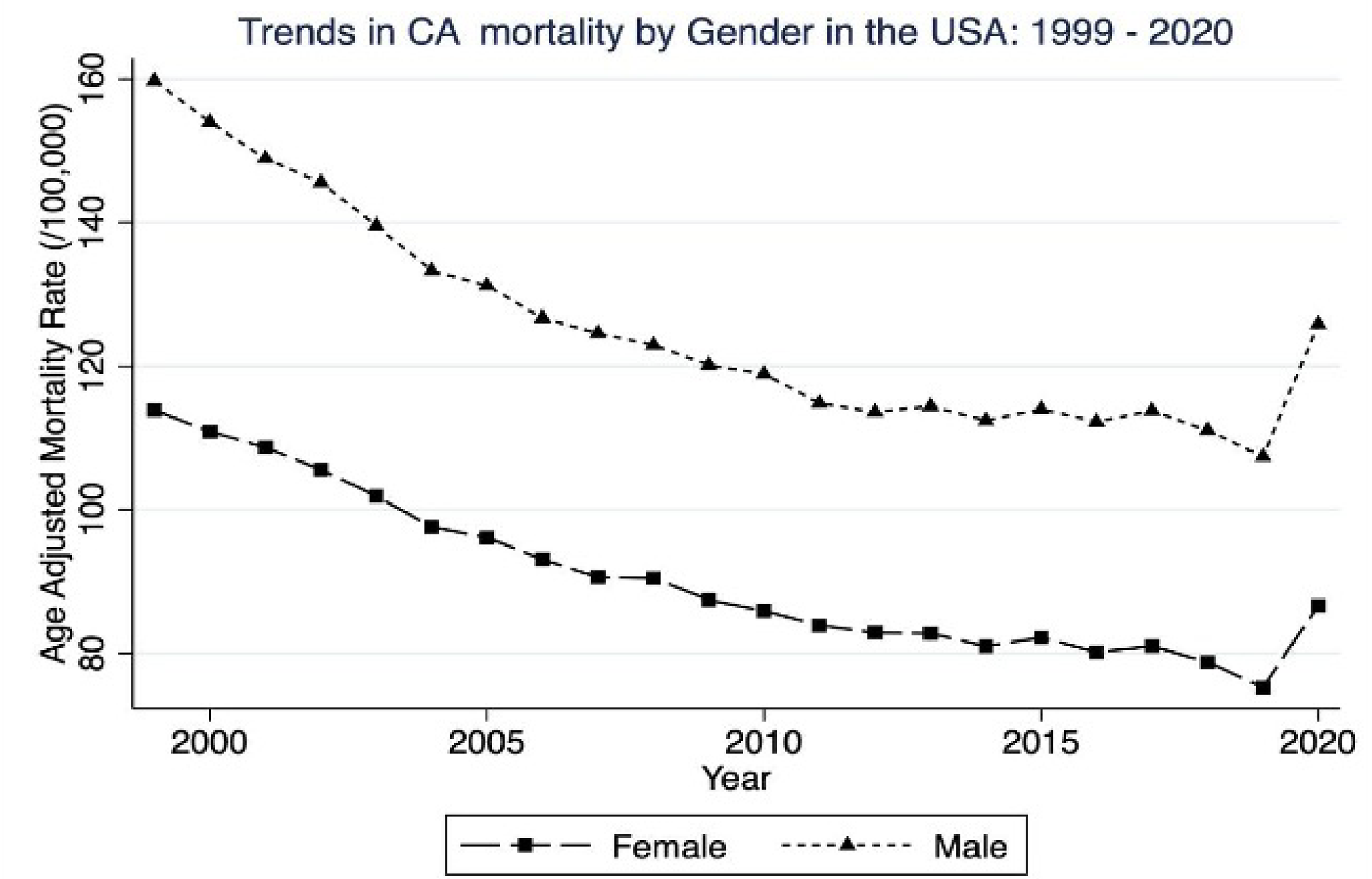
Trends in cardiac arrests related mortality by sex

The four geographic regions in the US showed similar trends in AAMR between 1999 and 2020. AAMR in Northeast decreased from 188.24 per 100,000 persons in 1999 to 117.61 per 100,000 persons in 2019, with an increase to 142.6 per 100,000 persons in 2020. AAMR in the Midwest decreased from 72.69 per 100,000 persons in 1999 to 62.59 per 100,000 persons in 2019, with an increase to 70.39 per 100,000 persons in 2020. AAMR in the Southern region decreased from 128.08 per 100,000 persons in 1999 to 75.78 per 100,000 persons in 2019, with an increase to 86.22 per 100,000 persons in 2020, and in the West, it decreased from 155.69 per 100,000 persons in 1999 to 115.03 per 100,000 persons in 2019, with an increase to 135.29 per 100,000 persons in 2020. (Figure 1c). We also noted significant differences within a region in regard to CA-MR. States with the highest AAMR included(per 100,000 persons): New York 254.83, Mississippi 218.3, Georgia 195.4, California 187.95, Nevada 178.94, Connecticut 169.7, Alabama 143.47, Hawaii 142.55, Nebraska 132.93, West Virginia 115.43, Massachusetts 105.07. (Figure 2). There was significant state-wide variation in the CA-MR over 22 years. The majority of the states showed an overall decrease in AAMR from 1999-2020 [Maine (−5.8), District of Columbia (−4.9), Hawaii (−5.2), West Virginia (−5.2), Nebraska (−4.1), Florida (−3.2), Georgia (−3), New York (−2.8), Vermont (−2.5), Missouri (−2.4), Tennessee (−2.3), Kentucky (−1.9), Connecticut (−1.8), North Carolina (−1.8), Texas (−1.7), New Mexico (−1.5), Ohio (−1.1), California (−1), Pennsylvania (−0.8)]. However, certain states had an overall increase in AAMR [New Jersey (3.7), Arkansas (2.8), Oregon (2.4), Illinois (1.9), Minnesota (1.4), Michigan (1.3)]. (Figure 3). Data regarding AAMR across all races, genders, states, and regions, as well as the Joinpoint analyses, are presented in Supplement files.

**Figure 1c:**
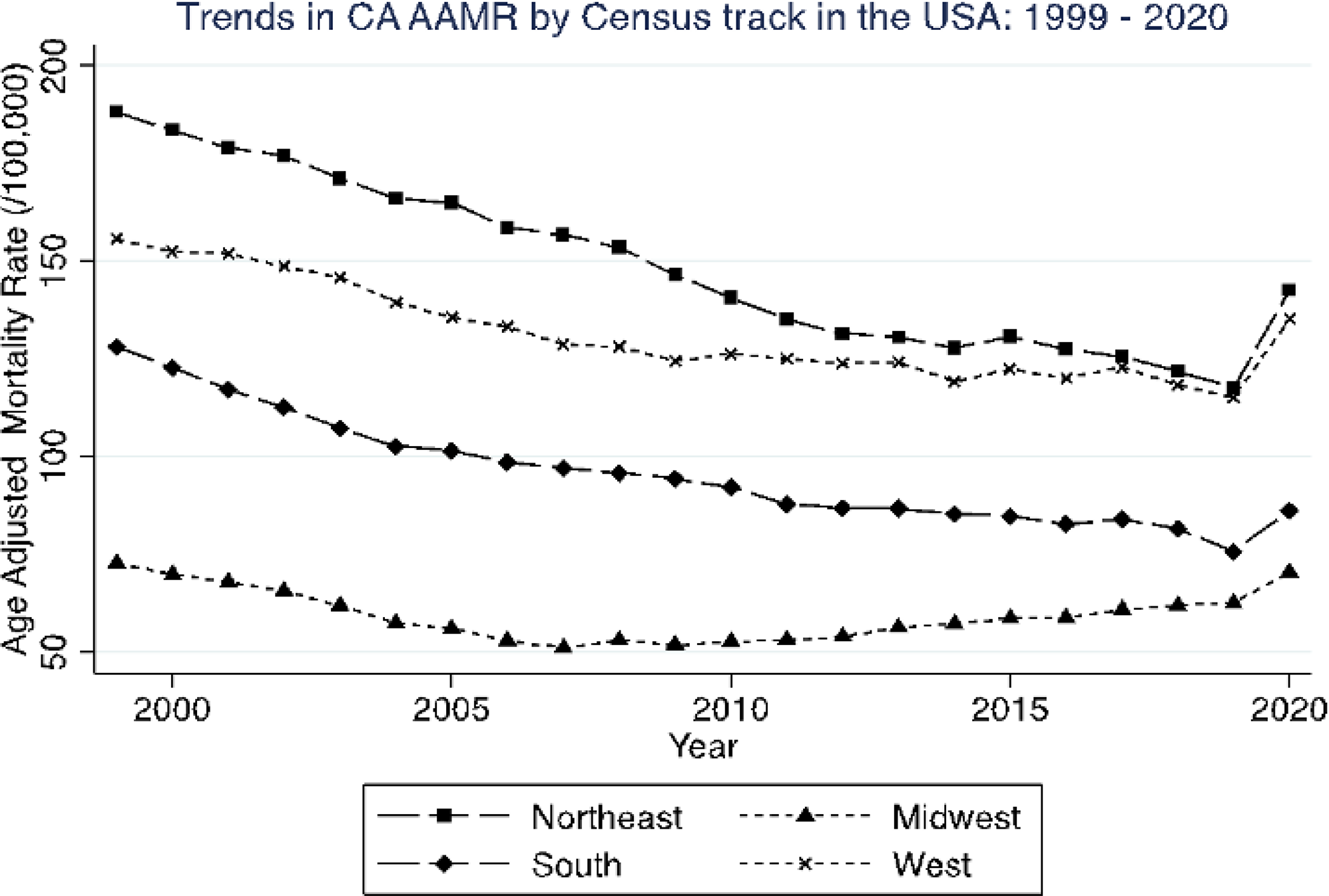
Trends in cardiac arrests related mortality by US regions

**Figure 2:**
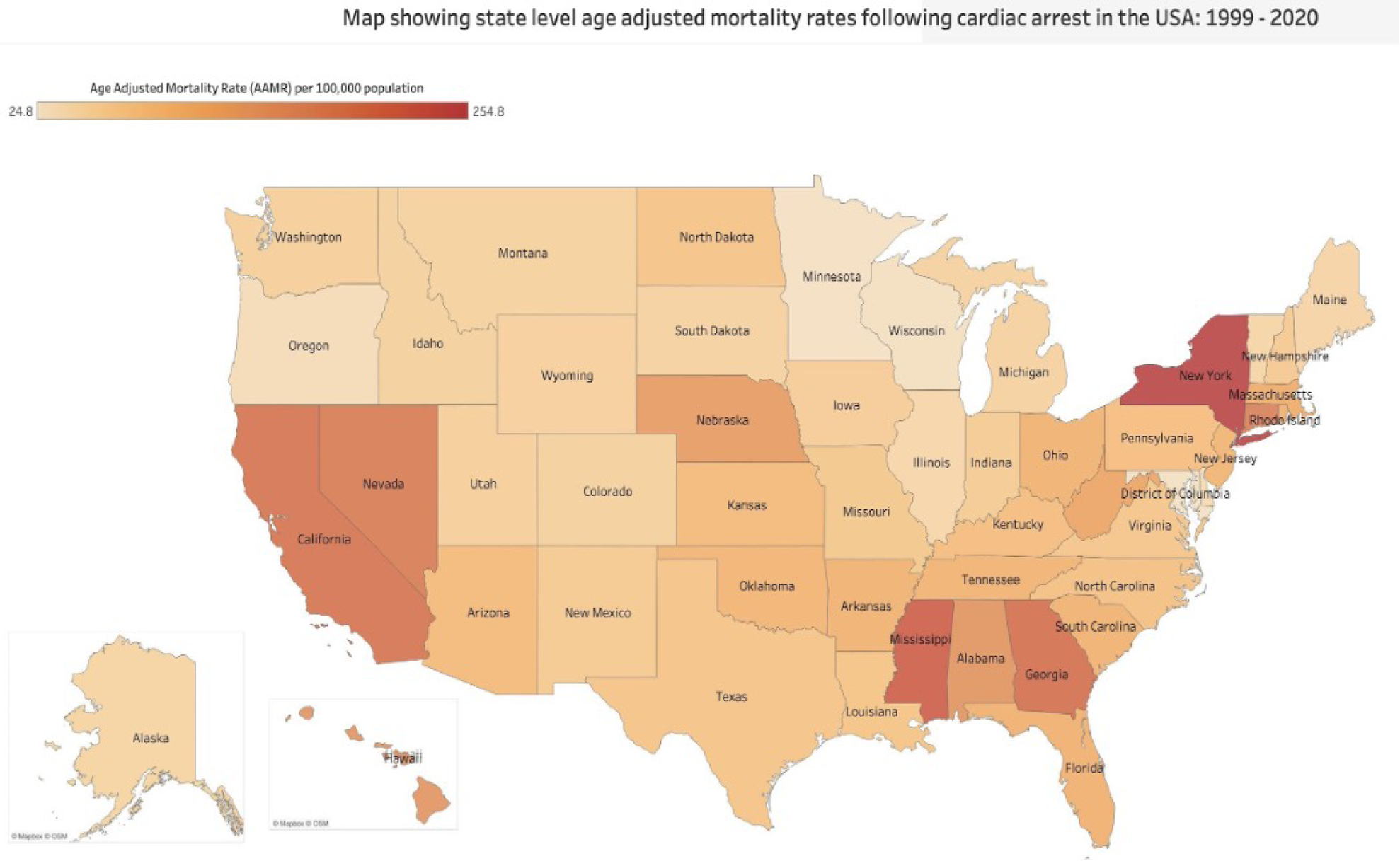
Map showing state-level cardiac arrests related age-adjusted mortality rates in the US.

**Figure 3:**
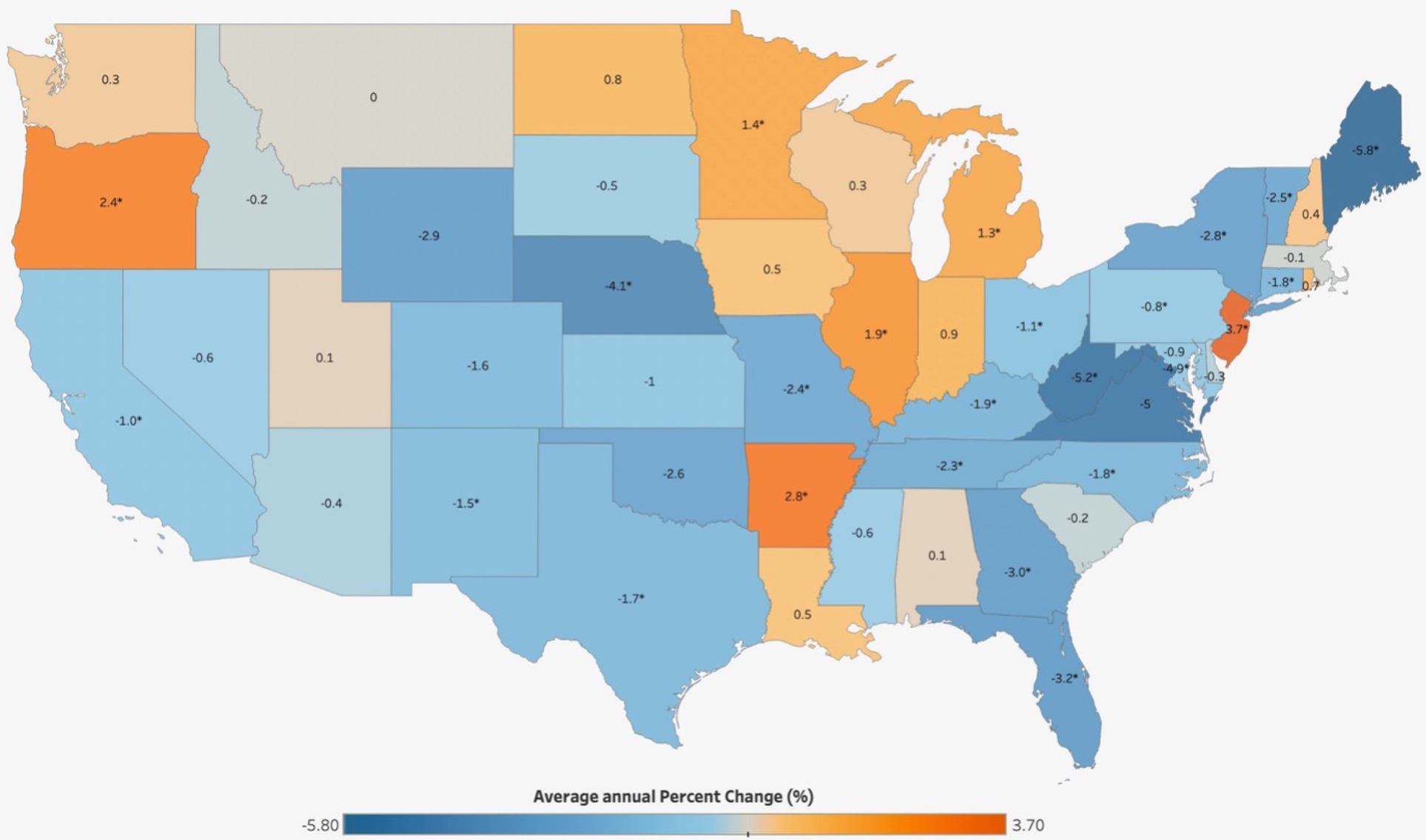
Map showing state-wise AAMR trend over 22 years.

## Discussion

We analyzed over 20 years of data using the CDC wonder database to assess the age-adjusted mortality rate among patients with cardiac arrest from 1999 to 2020 in the U.S. The principal findings in our study include:

1. There was an overall reduction in the age-adjusted mortality rates due to cardiac arrest. This decrease in mortality rate was consistent across all genders, races, and US regions.
2. Although there was a significant decrease in the mortality rates over time in the four US regions, a state-level variation was noted, with certain states showing an increase in mortality rates.
3. In the year 2020, a reversal in trend with an increase in the rates of cardiac arrest across all demographic subgroups was noted, likely secondary to the COVID-19 pandemic.

We report a favorable downward trend in CA-MR during the study period in the overall population and across age groups, various ethnic groups, and different regions. The downtrend in CA-related deaths could be from multiple factors. First, the incidence of myocardial infarctions (MI) has trended lower in recent years^19^. The decline in AMI was due to the dramatic decrease in the incidence of ST elevation MI, while the incidence of Non-ST Elevation MI may have increased. ST elevation MI is associated with significant scar burden compared to Non-ST Elevation MI.^20^ The time-sensitive care and interventions in AMI management also play a significant role. Myocardial scar serves as a typical substrate for ventricular arrhythmia leading to cardiac arrest. The decline in AMI is largely due to improved preventive care with increasing use of statins and modification of cardiovascular risk factors. The other etiologies for CA, such as channelopathies and non-ischemic cardiomyopathies, are unlikely to have seen a decline as they have a genetic component. Second, improvements in the chain of survival with high-quality CPR and also bystander CPR could also contribute to a decline in deaths during the time period. Indeed, Chan et al reported improved survival in OHCA in an analysis from a prospective registry from the US^21^. The improved survival was attributed to both improved pre-hospital and in-hospital survival. Third, newer American heart association (AHA) resuscitation guidelines focusing on in-hospital cardiac arrest patients’ care to better manage and standardize advanced cardiovascular life support (ACLS). It has led to improved post-resuscitation care such as therapeutic hypothermia, early percutaneous coronary intervention, and advanced critical care support.

Consistent with prior studies, women had significantly lower CA-MR compared to men.^3^. The possible explanations for this difference are women have cardioprotective factors that could potentially favor lesser mortality rates from CA than men. Women also have a higher studied likelihood of return of spontaneous circulation compared to men, and at baseline females tend to develop coronary artery disease (CAD) 10-15 years later than men^22,23^. The overall lifetime risk of sudden cardiac death (SCD) for men is 10.9% for men while it is 2.8% for women, and while the incidence of SCD increases with age for both males and females, a 2:1 ratio has been shown to exist between males and females across all ages. Furthermore, the proportion of SCD due to CAD remains lower in women, with an estimated 45%–50% versus 80%–90% in men^24^. While women may differ in baseline risk factors for having a CA, gender disparities amongst bystander CPR rates continue to suggest that further improvement in basic life support educational materials and training can be made, such as with the incorporation of female models. Studies have further suggested that public concern about legal, and cultural consequences, and hesitation with applying CPR skills to female anatomy were common sex-specific concerns. ^25^ Studies across different countries such as Sweden, Denmark, Netherlands, Japan, and the United States have shown women are less likely than men to receive bystander CPR^24^. This is further supported by studies that show males have an increased likelihood of receiving bystander CPR than females in public locations, but when at home where lay responders are mostly family members, gender disparities for bystander CPR delivery were not seen^26^.

Previous studies analyzed the association of race and socioeconomic status with mortality from cardiac arrest.^11,14,27,28^ Racial differences in the incidence and outcomes of cardiac arrest are well described. ^11,28,29^ Our study extends the observations of those prior studies reporting higher mortality rates in the African-American race compared to the White race. Data from the CPR Chicago project shows a higher incidence of CA and poor rates of survival in Black patients compared to White patients.^30^ The study reported that Black patients were less likely to have witnessed cardiac arrest, bystander-initiated CPR, or an initial shockable rhythm or to be admitted to the hospital. The odds of survival are higher for witnessed CA as there is a higher likelihood of early recognition and bystander CPR^31^. There is long-standing and unequivocal evidence of low rates of bystander CPR in Black patients and even adjusted for neighborhood income and location of CA, partly attributable to explicit and implicit bias^28^. Similar rates of limited bystander CPR initiation have also been seen amongst Latino and non-English speaking neighborhoods ^32^. Multiple studies have also shown lower rates of shockable rhythm in Black patients^11,28^. The relative contribution of differences in underlying comorbidities, genetic predisposition, or degeneration of shockable rhythm due to delay in intervention to the identified disparities remains to be seen. Further compounding the racial disparities is the allocation of resources in the Black/Hispanic predominant neighborhoods. Previous studies have shown a correlation between counties with higher African American and Hispanic residents and lower rates of CPR training^33,34^. Lower county-level rates of CPR training may, in part, contribute to lower rates of bystander CPR, lower rates of OHCA survival, and increased mortality from CA. Education about the recognition of cardiac arrest and population-based CPR training are community-level interventions that improve outcomes of CA, irrespective of race^35^. However, despite an improved rate of bystander interventions with community-level initiatives, Moeller et al reported improved survival only in White patients and not Black patients^32^. The entire chain of survival needs to be examined to identify the modifiable factors both at the prehospital level as well as the intra-hospital level to resolve racial disparities.

The Northeast had a higher CA-MR followed by the West, South, and Midwest. An interesting finding noted was that the AAMR dropped in the Midwest till 2007 followed by an increase in MR which is different from other regions as they showed a uniform decrease in the AAMR over 20 years. AAMR went up in 2020 across all regions except the South which could possibly be related to the delayed COVID outbreak in the Southern regions^36^. There were significant regional differences in the burden of CA-MR and states which were in the upper 90^th^ percentile of cardiac arrest were New York, Mississippi, Georgia, California, Nevada, Connecticut, Alabama, Hawaii, Nebraska, West Virginia, and Massachusetts. The potential explanation as to why regions with high population density, such as in the Northeast and West have higher CA-MR could be due to longer response time and delay in CPR initiation, lack of early defibrillation, and longer transportation time to the hospital^35^. Not only EMS operations but factors beyond the field such as healthcare setup/provider distribution plays a key component. Baseline patient’s cardiovascular health risk factors such as obesity, smoking, diet, diabetes, and substance use also show substantial variability across the United States and can contribute to CA-MR^37^. Limited resource availability, from either geographic spacing or population density, baseline variability in cardiovascular risk factors, and variation in CPR education rates are areas that require further public health investigation to mitigate CA care gaps. Following a steady decline in CA-MR from 1999 to 2019, a reversal in trend with an increase in AAMR was seen in 2020, consistent across sex, racial, and geographic subgroups. A similar reversal of the trend with increased mortality due to cardiovascular diseases in 2020 has been reported^38^. This trend correlates with the start of the COVID-19 pandemic and the resultant unprecedented strain on healthcare and emergency response systems. Chan et al. reported an increased incidence of CA during the pandemic, with a significant increase in counties most affected by COVID-19. Compared to the pre-COVID-19 pandemic phase, the pandemic period was associated with a lower likelihood of sustained ROSC after an OHCA, a higher likelihood of termination of CPR in the field, lower survival rates to discharge from CA, and less favorable neurological recovery compared to the pre-COVID-19 period^39–41^.

### Limitations

Even though the CDC WONDER is a large database, there are some limitations to the study due to shortcomings of the database. The dataset is obtained from death certificates and relies on the accuracy of data in the death certificate. The data is obtained exclusively from death reports and does not include information about co-morbidities, prior medical and surgical history, medications, vital signs, laboratory information, treatment, and information about CPR and AED utilization. Causes of death are listed using ICD coding which are susceptible to coding errors. We performed a retrospective cross-sectional analysis, so we were not able to prove causal relationships between county-level characteristics and mortality. Individual risk factors cannot be examined, so we performed crude estimates of AAMR. The database provides us with aggregated data and not individual-level data which limits our ability to perform adjusted analyses. To control these limitations, we performed a subgroup analysis looking at demographic and population-level variables. A considerable number of OHCA can go unassessed, so the likely cardiac arrest burden is significantly higher. Cardiac arrest in our cohort includes both out-of-hospital and in-hospital cardiac arrest. The etiology of cardiac arrest– traumatic vs. non-traumatic cannot be discerned.

## Conclusion

We report improvements in AAMR due to cardiac arrest in the overall US population and across most demographic groups with higher mortality rates in the Black population. A multi-dimensional approach with an emphasis on social determinants of health could allow for targeted interventions to address the increased AAMR due to CA in vulnerable groups.

## Data Availability

Data derived from public domain resources

## Abbreviations

CA: Cardiac arrest
CA-MR: Cardiac arrest related age adjusted mortality rate
CDC: Centers for Disease Control and Prevention
WONDER: Wide-Ranging Online Data for Epidemiologic Research
SVI: Social vulnerability index
NCHS: National Center for Health Statistics Urban-Rural Classification Scheme
SDOH: Social determinants of health
ICD: International Classification of Diseases
AAMR: Age-adjusted mortality rates
APC: Annual percentage change
AAPC: Average annual percentage change
CI: confidence interval
AHA: American heart association
CPR: Cardiopulmonary resuscitation
ACLS: Advanced cardiovascular life support
AED: Automated external defibrillator
EMS: Emergency medical services

